# Improving 10-year cardiovascular risk prediction in patients with type 2 diabetes with metabolomics

**DOI:** 10.1101/2024.04.16.24305917

**Authors:** Ruijie Xie, Teresa Seum, Sha Sha, Kira Trares, Bernd Holleczek, Hermann Brenner, Ben Schöttker

**Affiliations:** Division of Clinical Epidemiology and Aging Research, German Cancer Research Center, Im Neuenheimer Feld 581, 69120 Heidelberg, Germany; Faculty of Medicine, University of Heidelberg, 69115 Heidelberg, Germany; Saarland Cancer Registry, Neugeländstraße 9, 66117 Saarbrücken, Germany

**Keywords:** Type 2 diabetes, metabolomics, cardiovascular risk, prediction model

## Abstract

**Background and Aims:** To evaluate the potential of improved prediction of the 10-year risk of major adverse cardiovascular events (MACE) in patients with type 2 diabetes by adding metabolomic biomarkers to the SCORE2-Diabetes model.

**Methods:** Data from 10,257 and 1,039 patients with type 2 diabetes from the UK Biobank (UKB) and the German ESTHER cohort, respectively, were used for model derivation, internal and external validation. A total of 249 metabolites were measured with nuclear magnetic resonance (NMR) spectroscopy. LASSO regression with bootstrapping was used to identify metabolites in sex-specific analyses and the predictive performance of metabolites added to the SCORE2-Diabetes model was primarily evaluated with Harrell’s C-index.

**Results:** Seven metabolomic biomarkers were selected by LASSO regression for enhanced MACE risk prediction (three for both sexes, three male- and one female-specific metabolite(s)). Especially albumin and the omega-3-fatty-acids-to-total-fatty-acids-percentage among males and lactate among females improved the C-index. In internal validation with 30% of the UKB, adding the selected metabolites to the SCORE2-Diabetes model increased the C-index statistically significantly (*P*=0.034) from 0.660 to 0.680 in the total sample. In external validation with ESTHER, the C-index increase was higher (+0.041) and remained statistically significant (*P*=0.015).

**Conclusions:** Incorporating seven metabolomic biomarkers in the SCORE2-Diabetes model enhanced its ability to predict MACE in patients with type 2 diabetes. Given the latest cost reduction and standardization efforts, NMR metabolomics has the potential for translation into the clinical routine.

## Introduction

Cardiovascular diseases (CVDs) remain the leading cause of death in Europe, contributing to over 60 million potential life-years lost annually (1). In particular, individuals with type 2 diabetes have an increased incidence of CVD, with a risk profile that differs significantly from the general population (2). In light of this, identifying high-risk subgroups offers potentials for optimizing healthcare resource allocation and initiating preventive strategies against CVD events (3).

To further refine this cardiovascular risk evaluation specifically for people with type 2 diabetes, the SCORE2-Diabetes model became available in 2023 (4). It was designed to stratify individuals with type 2 diabetes into distinct risk categories based on their 10-year risk of major adverse cardiovascular events (MACE). Despite the advancement by the SCORE2-Diabetes model, a need for an enhanced precision in risk evaluation for this susceptible population still persists (5).

Metabolomics offers an unrivaled perspective through analyzing minuscule molecular metabolites within specific tissues or biofluids. It resonates with the genetic, environmental, and pathophysiological alterations characterizing the progression of type 2 diabetes and CVD (6). Harnessing such insights holds promise to enhance the predictive capacities of traditional prediction models by unveiling metabolites intrinsically associated with CVD risk.

Although several studies have examined the role of metabolites in improving the prediction of MACE in patients with type 2 diabetes, most were limited by small sample size, short follow-up durations, and a lack of validation in external cohorts (7–10). Additionally, emerging evidence emphasizes that there are sex differences in the associations of some metabolites with MACE but past studies have typically used sex as a covariate rather than selecting metabolomic biomarkers based on sex specificity (11; 12).

The present study aimed to leverage data from two large European cohorts to select metabolomic biomarkers based on a sex-specific selection and to assess whether integration of these metabolomic biomarkers improve the predictive power of the SCORE2-Diabetes model for a 10-year cardiovascular risk assessment.

## Methods

### Study design and population

The UK Biobank (UKB), a large prospective cohort in the United Kingdom, aims to bolster knowledge, preventive measures, diagnostic modalities, and therapeutic approaches across diverse disease spectra (13). In brief, between 13 March 2006 and 1 October 2010, over half million participants aged 37-73 were enrolled across England, Scotland, and Wales.

The ESTHER study is a large population-based cohort study conducted in Saarland, Germany. Participants aged between 50 and 75 years were recruited during a regular health check by local general practitioners (GP) between 1 July 2000 and 30 June 2002. After that, participants were followed up every two to three years. The study effectively collected data on 9,940 participants (14; 15).

### Metabolites quantification

Nightingale Health’s high-throughput nuclear magnetic resonance (NMR) metabolomics platform performed the metabolomic profiling on 274,353 baseline plasma samples from randomly selected UKB participants, alongside all available 8,308 baseline serum samples from the ESTHER cohort with sufficient blood sample quality (16). This process enabled the quantification of 250 metabolomic biomarkers, including lipids, fatty acids, amino acids, ketone bodies, and other essential low-molecular-weight markers (17). However, glycerol was excluded because it could not be measured in most of the participants of both cohorts, leaving n=249 metabolites for the analyses. The nomenclature and completeness of these metabolites are shown in **Supplemental Table S1**.

### Variables of the SCORE2-Diabetes model

The SCORE2-Diabetes is a sex-specific competing risk-adjusted algorithm recently developed and validated by the European Society of Cardiology, designed for adults with type 2 diabetes aged 40 to 69 years, includes age, sex, systolic blood pressure (SBP), smoking status, age at diabetes diagnosis, HbA_1c_, and the estimated glomerular filtration rate (eGFR) (4).

Information on demographic characteristics, lifestyle factors and medical history, including age, sex, age at diabetes diagnosis and smoking status (current or non-current) was collected by standardized questionnaires in both cohorts. The SBP measurements were conducted by automated reading of the Omron digital blood pressure monitor at the left upper arm in UKB and from the physician’s medical conditions report of the health check-up in the ESTHER study. The laboratory methods used to measure HDL-C, total cholesterol, HbA1c and creatinine in both cohorts are shown in **Supplemental Table S2**. The eGFR was calculated from creatinine values using the CKD-Epi 2009 equation in both cohorts (18).

### Outcome assessment

The primary outcome was MACE, which included cardiovascular mortality, non-fatal myocardial infarction, and non-fatal stroke (see detailed definition in **Supplemental Table S3**). It was constructed in line with the endpoint definition used to derive the SCORE2-Diabetes model. The only exception was that the non-fatal stroke definition was broader in the ESTHER study because stroke subtypes cannot be distinguished in this cohort.

In the UKB cohort, non-fatal myocardial infarction and or non-fatal stroke were identified from primary care records or hospital episode statistics. The date and cause of death were determined by referring to death registries of the National Health Service Information Centre for participants in the UKB study from England and Wales, and the National Health Service Central Register of Scotland for those from Scotland.

As previously described (19), study participants from the ESTHER study reported on the occurrence of incident myocardial infarction and stroke in standardized questionnaires at 2-, 5, 8-, and 11-year follow-ups. Self-reported cases underwent validation by the dissemination of standardized questionnaires to the GPs of the study participants. The proportions of physician-validated incident myocardial infarction (MI) and stroke cases were 89% and 91%, respectively. A vital status inquiry was made at registration offices of residents, with death certificates of deceased individuals being procured from local health authorities.

The follow-up period extended until the first recorded non-fatal myocardial infarction, non-fatal stroke, death, or the end of the event registration period, which was capped at ten years (4).

### In- and exclusion criteria

We included n=10,257 and n=1,039 individuals with type 2 diabetes and measured metabolomics data from the UKB and ESTHER study, respectively (**Supplemental Figure S1**). Subjects with potential type 1 diabetes, ascertained by diabetes diagnosis before the 40^th^ birthday, were excluded. UKB participants outside the age range of 50-74 years were excluded to make the age distribution of two cohorts comparable. Furthermore, those with a history of myocardial infarction or stroke before baseline or with a missing date on these events of interest were excluded.

### Statistical analyses

#### General remarks

All analyses were conducted using R software, version 4.3.0 (R Foundation for Statistical Computing, Vienna, Austria), with statistical significance set at *P*-values<0.05 for two-sided tests. Missing values were imputed using the random forest imputation method, as implemented in the r-package *missForest* (version 1.5) (20). Most variables of the SCORE2-Diabetes model and the NMR metabolites were complete. **Supplemental Table S1** illustrates that the highest missing value rate was observed for HbA1c in the UKB at 5.1%. In the ESTHER study, aside from the age at diabetes diagnosis (39.6%), no other variable’s missing rate exceeded 10%.

#### Metabolites selection and model derivation

Initially, all metabolite concentrations underwent log-transformation to approximate normal distributions for analysis. These measures were then scaled to standard deviation units independently within each cohort. The UKB dataset was partitioned into a derivation set (70%) and an internal validation set (30%); the ESTHER study served as the external validation cohort. The derivation set was used for metabolite selection and model derivation.

The Least Absolute Shrinkage and Selection Operator (LASSO), a regularization technique adept at identifying strong predictors among high-dimensional and correlated predictors, was implemented via the r-package *glmnet* (version 4.1-7) (21). Ten-fold cross-validation was employed to determine the optimal tuning parameter λ for LASSO, based on the lowest model validation error. A bootstrap LASSO approach was then undertaken, involving the creation of 1000 derived sets with the LASSO procedure applied to each resampled dataset (22). Metabolites selected by LASSO in at least 95% of instances were designated as our metabolites of interest, a threshold adopted from a previous study (23) to enhance model generalization and mitigate overfitting. These selected metabolites were subsequently incorporated into the SCORE2-Diabetes model to construct new sex-specific competing risk models using the r-package *cmprsk* (version 2.2-11).

#### Model predictive performance validation

The predictive performance of the derived model was validated by using 30% of the UKB as the internal validation cohort and the ESTHER cohort as the external validation cohort. The incremental discrimination contributed by each metabolite was evaluated based on the internal validation results.

Discriminative ability was assessed using Harrell’s C-index, adjusted for competing risks (via the r-package *riskRegression*, version 2023.03.2) (24). Furthermore, the net reclassification index (NRI) and the integrated discrimination improvement (IDI) were evaluated for the final model (using the r-package *nricens*, version 1.6) (25). Pre-specified CVD risk categories (0– 10%, >10–20%, and >20%) were employed for the NRI to indicate the proportion of individuals correctly reclassified compared with the SCORE2-Diabetes model. Model calibration was examined by plotting observed MACE event rates against predicted event rates across deciles of absolute predicted risk.

#### Associations of selected metabolites with MACE

To elucidate the hazard ratios (HRs) and 95% confidence intervals (CIs) of selected metabolites (per one standard deviation increment) for enhanced 10-year MACE prediction in male and female patients with type 2 diabetes, metabolites were individually added to Cox proportional hazards regression models in the derivation cohort, the internal validation cohort, and the external validation cohort, respectively. These models were adjusted for SCORE2-Diabetes model variables, using the r-package *survival* (version 3.5-5). The Benjamini-Hochberg procedure was applied to control the false discovery rate (FDR) in this analysis.

## Results

### Baseline characteristics and MACE case numbers

**Table 1** summarizes the characteristics of the participants for all variables of the SCORE2-Diabetes model from the 10,257 and 1,039 included participants with type 2 diabetes from the UKB and ESTHER cohorts, respectively. The included UKB participants had an average age of 61.5 years (SD 5.2), with males constituting 59.9% of the cohort. Over a follow-up time limited to 10 years, 1,366 MACE events were recorded in the UKB. The ESTHER sample had a mean age of 64.0 years (SD 6.3), and 50.8% of participants were males. During the first 10 years of ESTHER follow-up, 175 MACE events were recorded.

**Table 1.** Baseline characteristics of selected participants with type 2 diabetes from the UK Biobank and ESTHER study.

| Baseline characteristics | UK Biobank (n=10,257) | ESTHER (n=1,039) |
| --- | --- | --- |
| Male sex, N (%) | 6,140 (59.9) | 528 (50.8) |
| Age (years), mean (SD) | 61.5 (5.2) | 64.0 (6.3) |
| Current smoker, N (%) | 1,024 (10.0) | 157 (15.7) |
| Systolic blood pressure (mmHg), mean (SD) | 144.8 (18.2) | 144.2 (19.5) |
| Total cholesterol (mmol/L), mean (SD) | 3.7 (0.9) | 5.6 (1.1) |
| HDL cholesterol (mmol/L), mean (SD) | 1.1 (0.3) | 1.3 (0.3) |
| Age at diabetes diagnosis (years), mean (SD) | 55.4 (6.4) | 55.6 (5.7) |
| HbA<sub>1c</sub> (mmol/mol), mean (SD) | 51.8 (12.8) | 53.1 (14.4) |
| eGFR (ml/min/1.73m<sup>2</sup>)<sup>a</sup>, mean (SD) | 87.7 (15.4) | 78.6 (17.7) |

<sup>a</sup> The eGFR was calculated with the CKD-EPI 2009 equation.
**Abbreviations:** eGFR, estimated Glomerular Filtration Rate; HbA<sub>1c</sub>, glycated hemoglobin; HDL, high-density lipoprotein; IQR, interquartile range; SD, standard deviation.

### Associations of metabolomic biomarkers with MACE

A total of 7 metabolites were selected to enhance MACE risk prediction in the SCORE2-diabetes model in the UKB cohort by LASSO analyses and bootstrapping with a frequency of more than 950 times in the 1000 replicate selection (**Supplemental Table S4**). Of these metabolites, three metabolites improved MACE risk prediction in both men and women (creatinine, albumin, and GlycA (glycoprotein acetyls)). Furthermore, lactate emerged as an additional predictive metabolite solely in women, while the three metabolites - acetate, VLDL-size (the average diameter for very low-density lipoprotein particles), and Omega-3-pct (the Omega-3 fatty acids to total fatty acids percentage) exhibited enhanced predictive capabilities exclusively in males.

Figure 1 shows the associations of the 7 selected metabolites with MACE in multivariate Cox regression models adjusted for SCORE2-Diabetes model variables separately in males and females in the derivation cohort. GlycA, and creatinine were positively, and omega-3-pct and albumin were inversely, statistically significantly associated with MACE at an FDR-adjusted *P*<0.05 level in both sexes. The results were generally consistent in the internal validation cohort but some results differed like for lactate for which results showed a sex difference in the internal validation study (**Supplemental Figure S2**) which was not seen in the derivation cohort (Figure 1) or the external validation cohort (**Supplemental Figure S3**). With the lower sample size of the ESTHER study, of the 7 metabolites, only albumin was statistically significantly associated with MACE in males.

**Figure 1.**
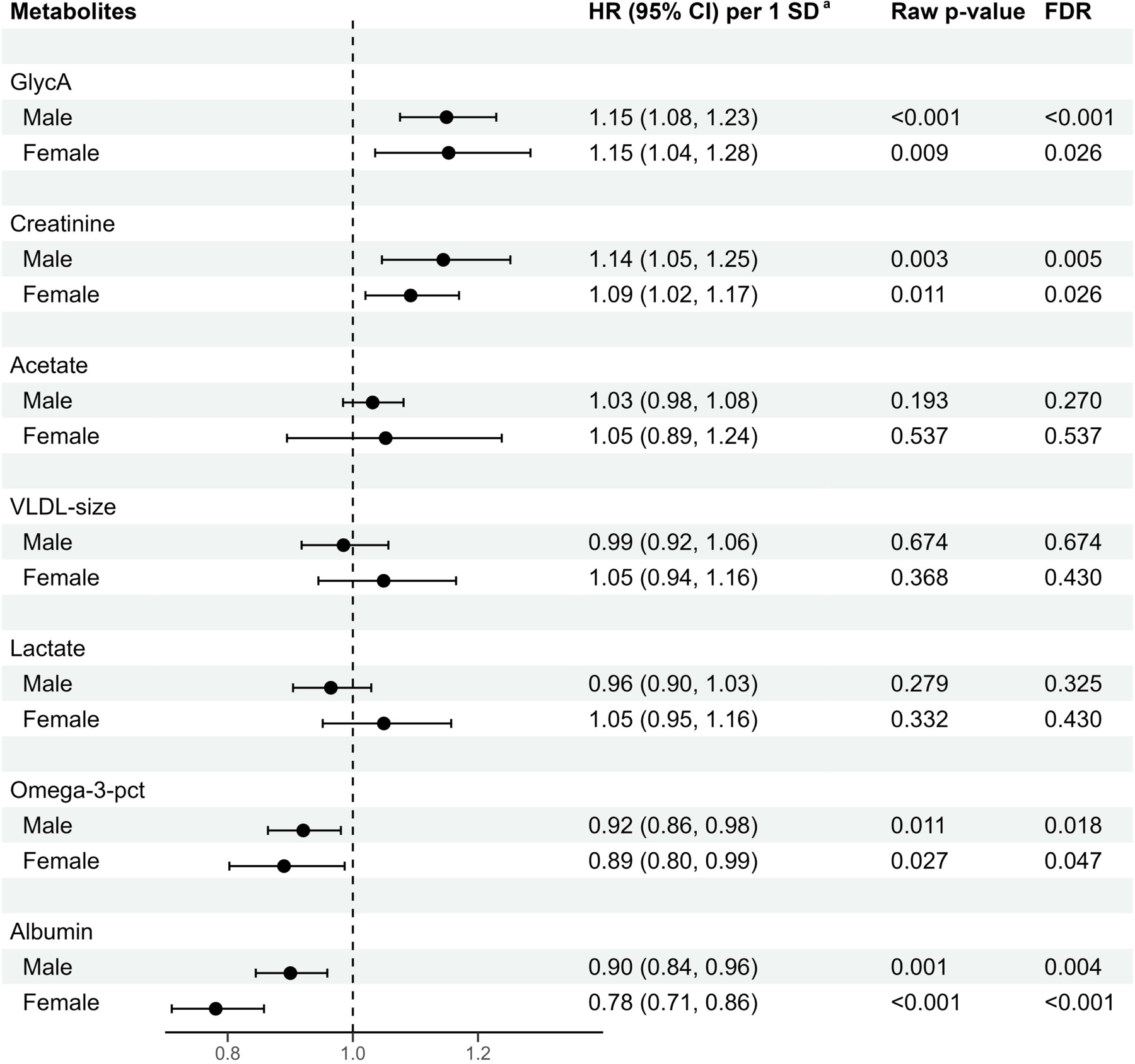
Associations between selected metabolites and major cardiovascular events across sexes in the derivation set (70% of UK biobank, N=7,180) **Abbreviations:** CI, confidence interval; FDR, false discovery rate; GlycA, glycoprotein acetyls; HR, hazard ratio; Omega-3-pct, Omega-3 fatty acids to total fatty acids percentage; SD, standard deviation; VLDL-size, average diameter for very-low-density lipoprotein particles. **^a^** Hazard ratios are expressed per 1 standard deviation of the respective metabolite concentration and are adjusted for age, systolic blood pressure, smoking status, diabetes age at diagnosis, glycated hemoglobin, and the estimated glomerular filtration rate. The standard deviations of creatinine, albumin, GlycA, acetate, omega-3-pct, VLDL-size, and lactate were 0.02 mmol/L, 3.60 mmol/L, 0.13 mmol/L, 0.04 mmol/L, 1.51%, 1.41 mmol/L, and 1.23 mmol/L, respectively.

### MACE risk prediction by metabolomic biomarkers

Figure 2 displays the incremental improvement in the C-statistic upon the sequential inclusion of the selected 7 metabolites into the models for males and females one by one in the internal validation set. Especially, the addition of albumin and Omega-3-pct significantly improved discrimination power in males. GlycA, creatinine, acetate, and VLDL-size showed notable but non-significant enhancements in males. In females, only lactate exhibited a notable, yet non-significant, C-index increase.

**Figure 2.**
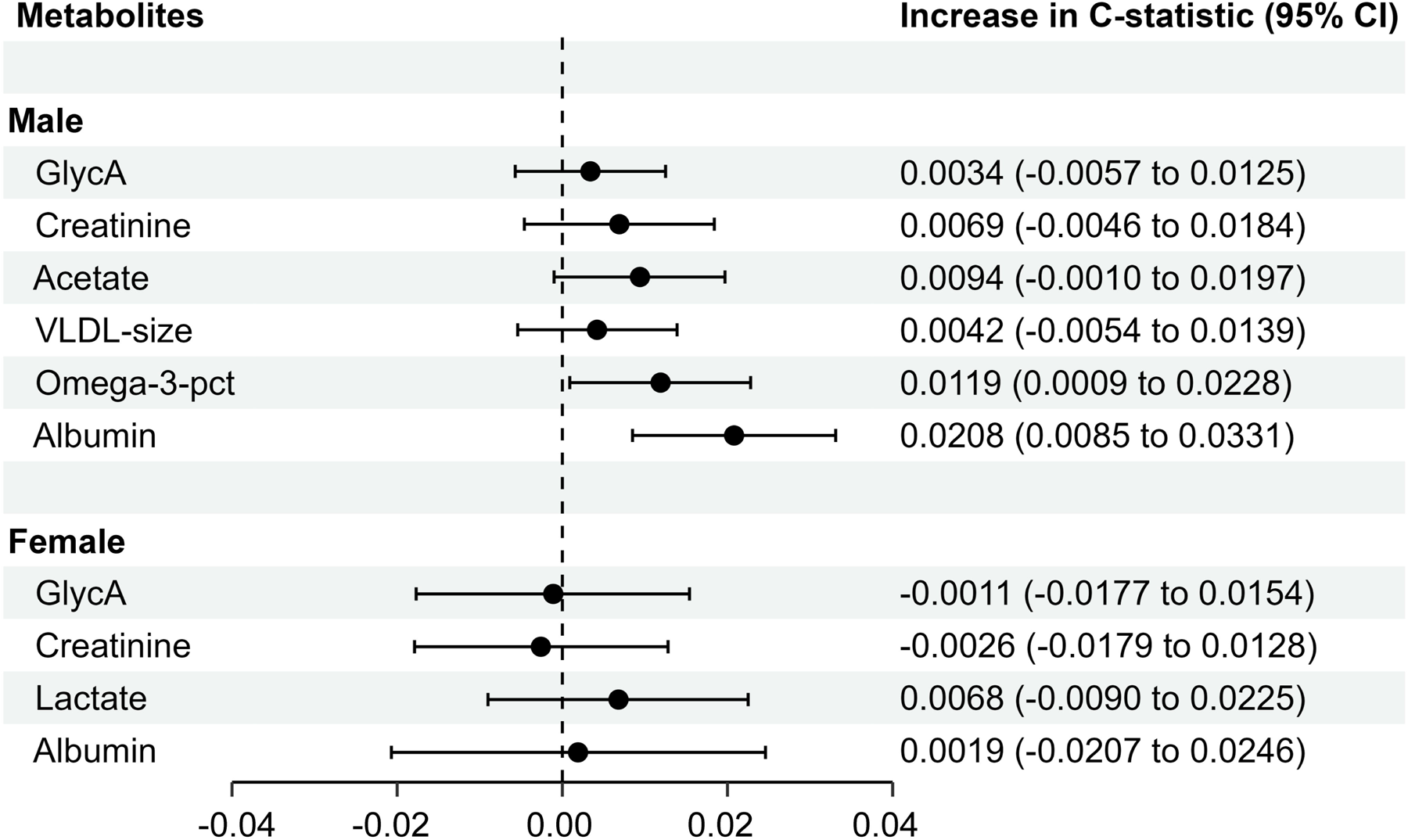
The incremental discrimination of each metabolite for the model after the selected metabolites were added to SCORE2-Diabetes separately in different sexes in the internal validation set (30% of UK Biobank, N=3,077) **Abbreviations:** CI, confidence interval; GlycA, glycoprotein acetyls; HR, hazard ratio; Omega-3-pct, Omega-3 fatty acids to total fatty acids percentage; VLDL-size, average diameter for very-low-density lipoprotein particles.

**Table 2** presents the predictive performance metrics of the SCORE2-Diabetes model for 10-year MACE risk prediction when used in combination with the 7 metabolomic biomarkers. The coefficients of all variables required for calculation of the extended risk model are shown in **Supplemental Table S4**. In the derivation set, by adding the selected metabolites, the C-index (95% CI) of the SCORE2-Diabetes model statistically significantly improved (*P*=0.008) from 0.662 (0.644-0.681) to 0.677 (0.659-0.696) in the total sample (**Table 2**). The C-index increase was more pronounced in males (+0.017) than females (+0.007), with statistical significance solely noted for males (*P* =0.020). Internal validation in the UKB yielded very similar results for the C-indices. Notably, an enhancement in risk reclassification statistics was primarily observed in female participants with a statistically significant IDI. Model calibration was comparably good for the SCORE2-Diabetes and the extended model in the internal validation set (**Supplemental Figure S4**).

**Table 2.** Metrics of the predictive performance of the SCORE2-Diabetes model for 10-year MACE risk without and with extension by metabolites.

| Metrics | Men | Women | Overall |
| --- | --- | --- | --- |
| **Derivation set (70% of UK Biobank)** |  |  |  |
| Total sample size / MACE cases | 4,298/691 | 2,882/249 | 7,180/940 |
| C-index (SCORE2-Diabetes) | 0.630 (0.606,0.653) | 0.673 (0.638,0.707) | 0.662 (0.644,0.681) |
| C-index (+Metabolites<sup>a,b</sup>) | 0.647 (0.624,0.670) | 0.680 (0.645, 0.714) | 0.677 (0.659,0.696) |
| P-values for C-index comparisons | **0.020** | 0.053 | **0.008** |
| **Internal validation set (30% of UK Biobank)** |  |  |  |
| Total sample size / MACE cases | 1,842/313 | 1,235/113 | 3,077/426 |
| C-index (SCORE2-Diabetes) | 0.632 (0.597, 0.668) | 0.670 (0.617, 0.722) | 0.660 (0.632, 0.688) |
| C-index (+Metabolites<sup>a,b</sup>) | 0.657 (0.623,0.692) | 0.679 (0.625,0.733) | 0.680 (0.652,0.708) |
| P-values for C-index comparisons | **0.005** | 0.716 | **0.034** |
| NRI<sup>c</sup> total (%) | 0.9 (-7.4, 9.3) | 7.0 (-8.8, 24.5) | 5.6 (-2.5, 13.8) |
| NRI<sup>c</sup> events (%) | 3.6 (-4.0, 7.5) | 2.3 (-14.0, 26.9) | 0.6 (-3.8, 7.9) |
| NRI<sup>c</sup> non-events (%) | -2.7 (-7.7, 3.8) | 4.7 (-4.5, 12.6) | 5.0 (-1.8, 8.8) |
| IDI | 0.000 (-0.015, 0.014) | **0.024 (0.002, 0.053)** | **0.013 (0.001, 0.026)** |
| **External validation set (ESTHER)** |  |  |  |
| Total sample size / MACE cases | 528/107 | 511/68 | 1,039/175 |
| C-index (SCORE2-Diabetes) | 0.632 (0.573,0.691) | 0.618 (0.545,0.691) | 0.612 (0.566,0.657) |
| C-index (+Metabolites<sup>a,b</sup>) | 0.663 (0.608, 0.719) | 0.648 (0.575, 0.721) | 0.653 (0.609, 0.697) |
| P-values for C-index comparisons | 0.178 | 0.241 | **0.015** |
| NRI<sup>c</sup> total (%) | 7.6 (-6.5, 16.0) | -6.9 (-23.7, 16.9) | 4.3 (-5.7, 12.9) |
| NRI<sup>c</sup> events (%) | **9.6 (0.9, 16.1)** | -3.0 (-16.0, 15.7) | 4.3 (-4.0, 10.5) |
| NRI<sup>c</sup> non-events (%) | -2.0 (-16.5, 10.2) | -3.9 (-17.4, 10.5) | 0.0 (-7.9, 8.7) |
| IDI | 0.004 (-0.025, 0.036) | 0.000 (-0.032, 0.042) | 0.010 (-0.007, 0.030) |

**Abbreviations:** IDI, integrated discrimination improvement; MACE, major cardiovascular events; NRI, net reclassification improvement.
<sup>a</sup> Metabolites that were included in the combined model for men were creatinine, albumin, GlycA, acetate, Omega-3-pct, and VLDL-size.
<sup>b</sup> Metabolites that were included in the combined model for women were lactate, creatinine, albumin, and GlycA.
<sup>c</sup> NRI calculated with the pre-specified 10-year MACE risk categories of 0–10%, >10–20%, and >20%.

In the external validation with the total ESTHER study, the model demonstrated an even greater improvement in discriminatory power compared to the UKB, with a likewise statistically significant result (the C-index increased from 0.612 to 0.653, *P*=0.015; **Table 2**). C-index increases were comparable for men (+0.031) and women (+0.030). Beyond these improvements, the reclassification metrics for males indicated that 9.6% of events were correctly reclassified by the extended model compared with the SCORE2-Diabetes model [NRI= 9.6% (0.9%, 16.1%)]. In males, the SCORE2-Diabetes model showed a good calibration, which was even better with the extended model (**Supplemental Figure S5**). In women, the SCORE2-Diabetes model was slightly better calibrated than the extended model. However, the MACE case number was quite low for female patients with type 2 diabetes in the ESTHER cohort (n=68), which should be taken into consideration when interpreting the figure.

## Discussion

Using large-scale data from two European cohorts of middle-aged and older patients with type 2 diabetes, we developed and validated a sex-specific competing risk algorithm that combines the SCORE2-Diabetes model with 7 metabolites to improve 10-year cardiovascular risk prediction. In internal and external validations, the C-index of the SCORE2-Diabetes model significantly increased by 0.020 and 0.041, respectively.

### Comparison with previous studies

There are a limited number of studies using metabolomics to predict cardiovascular risk in patients with type 2 diabetes. Three studies based on the ADVANCE trial (sample size: n=3,576-3,779, follow-up: 5 years) investigated the association of specific types of fatty acids, amino acids, and sphingolipids with cardiovascular events and demonstrated the predictive value of these metabolites for cardiovascular risk in patients with type 2 diabetes (9; 10; 26). However, the enhancement of the predictive ability of the original models by these metabolomic biomarkers was relatively modest, with improvements in the C-statistic ranging between 0.005 and 0.020. This limited enhancement could be attributed to the investigation focusing only on a selection of a few metabolites. In contrast, the Edinburgh Type 2 Diabetes Study (sample size: n=1,066, follow-up: 10 years) used a broader metabolomics assay with 228 metabolites (23). By integrating 12 selected metabolites into the reference model, there was a notable increase in the C-statistic by 0.019; which was comparable to our findings. However, the models derived from the ADVANCE trial and the Edinburgh Type 2 Diabetes Study lacked external validation and their metabolomics measurements were not conducted by a commercial service provider. This limits the application and dissemination of their metabolomics-based risk models in clinical practice (27).

A growing body of evidence emphasizes that the associations of some metabolites with MACE differ in men and women, and deriving sex-specific risk models is a potential way to enhance the predictive power (28–30). Sex differences in the association with CVD risk of metabolomic biomarkers are biologically possible due to differences in sex hormones, sex chromosomes, and lifestyle differences (31–33). For example, 141 out of 179 lipid species were found to differ significantly between males and females in the Finnish GeneRISK cohort (28). However, sex-specific analyses are much more demanding in terms of sample size, which might be the reason why - to our knowledge - there are currently no other studies with diabetic patients, which derived sex-specific, metabolomics-based cardiovascular risk prediction models.

### Biological mechanisms of selected metabolites

Three metabolites (creatinine, albumin, and GlycA) were chosen to enhance MACE risk prediction for both sexes, with lactate additionally selected for women, and acetate, VLDL-size, and Omega-3-pct specifically selected for men. Creatinine and albumin are indicative of renal function. Patients with moderate to severe chronic kidney disease have an increased MACE risk (3; 34). GlycA reflects the glycosylation status of acute phase proteins and thus is a pro-inflammatory biomarker. Inflammatory processes are crucial in the development and progression of cardiovascular diseases (35). Lactate, a key byproduct of glycolysis, is often elevated during metabolic stress. Elevated lactate levels are associated with inflammation, endothelial dysfunction, and oxidative stress (36). Acetate is the most abundant endogenously produced short-chain fatty acid and has been found to be associated with weight loss, enhanced insulin sensitivity, and a reduced cardiovascular risk reduction (37). The size of VLDL particles is an independent predictor of atherosclerosis beyond traditional lipid measurements, highlighting its importance in cardiovascular risk stratification in type 2 diabetes (38). Omega-3 fatty acids are known for their beneficial effects on endothelial function and their role in reducing systemic inflammation (39). If associations of these biomarkers with MACE were not in the expected direction (e.g. for lactate), this might be explained by the comprehensive adjustment for the variables of the SCORE2-Diabetes model.

### Strengths and limitations

In terms of strengths, our study stands out for its large sample size (n = 10,257 for UKB, n = 1,039 for ESTHER), making it the largest metabolomics study focused on predicting the cardiovascular risk in individuals with type 2 diabetes so far. Additionally, this study is pioneering in selecting metabolomic biomarkers for cardiovascular risk prediction separately by sex and using an external validation cohort. Furthermore, model calibration was performed for a low risk (UK) and intermediate cardiovascular risk region (Germany).

However, our study has certain limitations. The ESTHER study may not be the ideal external validation cohort for our model due to its differences from the UKB, including a higher mean age (64 vs. 61.5 years), a higher proportion of fasting of at least 4 hours prior to blood sample donation (89.1% vs. 23.7%), which can be relevant for some metabolic biomarkers, and the differences in the ascertainment of non-fatal strokes, which did not allow for the exclusion of hemorrhagic strokes, as well as the regional difference of cardiovascular risk (low in UK and intermediate in Germany). These differences could have negatively influenced the results of the external validation. However, despite these cohort differences, the overall consistency in risk prediction results between the two cohorts provides support for the robustness of the derived model. Another limitation was this study may be generalizable to low-to-intermediate risk regions in Europe, larger validation cohorts are needed, which should originate not only from low to intermediate risk regions in Europe but shall also validate the new risk model in populations from high and very high-risk regions.

### Potential clinical utility of the derived model

The integration of 7 selected metabolomic biomarkers in the SCORE2-Diabetes model has led to a significant improvement in the discrimination of 10-year cardiovascular risk in patients with type 2 diabetes. This augmentation enhances the predictive power of the model without compromising the convenience offered by the conventional model, which requires only a brief physical examination, interview, and blood sample analysis.

While incorporating metabolomics into clinical practice does incur costs, it is important to note that the NMR method used for metabolomic analysis is generally more cost-effective and standardized compared to the LC-MS/MS (Liquid Chromatography-Mass Spectrometry/Mass Spectrometry) method (40). Additionally, commercial companies like Nightingale Health are available to handle large sample sizes in a short time, making this approach feasible for widespread clinical application.

However, before recommending routine measurement in clinical practice, it is essential to consider country-specific validation of the model. Currently, our algorithm is primarily validated for the UK and Germany, and extending this validation to other regions is crucial for broader applicability. The differences in cardiovascular risk profiles and healthcare systems across countries necessitate such validation to ensure the effectiveness and relevance of the model in diverse settings.

## Conclusions

This investigation showed for the first time with an external validation cohort, that metabolites can enhance an established MACE prediction model for patients with type 2 diabetes. As metabolomic analyses became standardized and affordable by the NMR technology in recent years, these measurements have a translation potential for clinical routine. The developed and validated SCORE2-Diabetes model extended by 7 metabolites can be used for an improved cardiovascular risk stratification, which is needed for personalized cardiovascular prevention measures in patients with type 2 diabetes.

## Data Availability

Data from ESTHER is available upon reasonable request that is compatible with participants' informed consent. Data from the UK Biobank (https://www.ukbiobank.ac.uk/) is available to bona fide researchers on application.

## List of abbreviations

CI: confidence intervals
CVD: cardiovascular disease
eGFR: estimated glomerular filtration rate
ESTHER: Epidemiologische Studie zu Chancen der Verhütung, Früherkennung und optimierten Therapie chronischer Erkrankungen in der älteren Bevölkerung
FDR: false discovery rat
GlycA: glycoprotein acetyls
GP: general practitioner
HR: hazard ratio; IDI, integrated discrimination improvement
LASSO: least absolute shrinkage and selection operator
LC-MS/MS: Liquid Chromatography-Mass Spectrometry/Mass Spectrometry
MACE: major adverse cardiovascular events
MI: myocardial infarction
NMR: nuclear magnetic resonance
NRI: net reclassification index; Omega-3-pct, the Omega-3 fatty acids percentage of total fatty acids
SBP: systolic blood pressure
SD: standard deviation
UKB: UK Biobank
VLDL-size: the average diameter for very low-density lipoprotein particles.

## Acknowledgements

We would like to thank all participants of the ESTHER and UK Biobank cohort as well as the GPs of the ESTHER study and the staff of the UK Biobank assessment centers for their contribution to the studies this research is based on. Part of this research was conducted using the UK Biobank Resource under application 101633.

## Article Information

### Author’s Contributions

H.B. designed and led the ESTHER cohort. R.X. and B.S. generated the idea for the study and formulated the analytical plan. R.X. did the data analyses and drafted the manuscript. B.S. revised it. All authors contributed valuable intellectual content to the discussion. The corresponding author attests that all listed authors meet authorship criteria and that no others meeting the criteria have been omitted. H.B. and B.S. had full access to ESTHER data. R.X. and B.S. had full access to UK Biobank data used for this study. R.X. and B.S. are the guarantors of the manuscript and accepts full responsibility for the work and/or the conduct of the study.

### Prior Presentation

Not applicable.

---

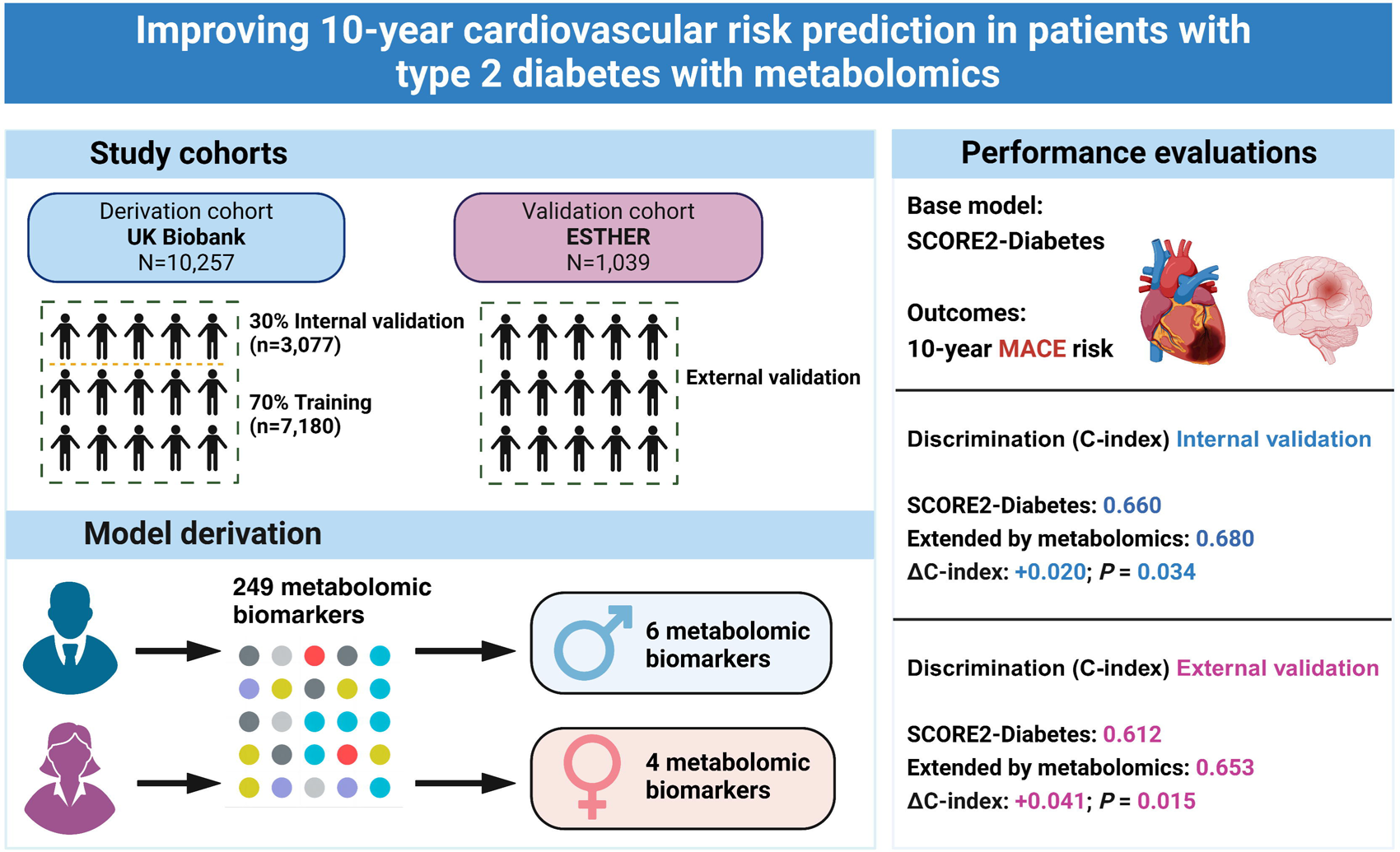

