## Supplemental Tables and Figures for "Improving 10-year cardiovascular risk prediction in patients with type 2 diabetes with metabolomics"

**Table of contents**

**[Table S1.](#_Toc18977)** [Completeness of baseline variables among the included study population from the UK Biobank and the ESTHER study before multiple imputation	2](#_Toc18977)

**[Table S2.](#_Toc29371)** [Laboratory methods applied in the UK Biobank (UKB) and ESTHER study	7](#_Toc29371)

**[Table S3.](#_Toc5167)** [Definition of endpoint major cardiovascular event (MACE)	8](#_Toc5167)

**[Table S4.](#_Toc1402)** [ß-coefficients of the variables of the SCORE2-Diabetes model extended by 7 metabolites for 10-year prediction of major cardiovascular events in the derivation set of the UK Biobank	9](#_Toc1402)

**[Figure S1.](#_Toc31318)** [Flow chart for participant inclusion and exclusion	10](#_Toc31318)

**[Figure S2.](#_Toc9342)** [Associations between selected metabolites and major cardiovascular events across sexes in the internal validation cohort (30% of UK Biobank, N=3,077)	11](#_Toc9342)

**[Figure S3.](#_Toc8962)** [Associations between selected metabolites and major cardiovascular events across sexes in the external validation cohort (ESTHER study, N=1,039)	12](#_Toc8962)

**[Figure S4.](#_Toc10994)** [Calibration curves of the SCORE2-Diabetes model and its combination with the metabolites for 10-year cardiovascular risk prediction in the internal validation set (30% of UK Biobank, N=3,077)	13](#_Toc10994)

**[Figure S5.](#_Toc22875)** [Calibration curves of the SCORE2-Diabetes model and its combination with the metabolites for 10-year cardiovascular risk prediction in the external validation (ESTHER study, N=1,039)	14](#_Toc22875)

**Table S1.** Completeness of baseline variables among the included study population from the UK Biobank and the ESTHER study before multiple imputation

| **Variable** | **UK Biobank  n (%)** | **ESTHER  (n (%)** |
| --- | --- | --- |
| age | 10257 (100.0) | 1039 (100.0) |
| sex | 10257 (100.0) | 1039 (100.0) |
| Systolic blood pressure | 10227 (99.7) | 1020 (98.2) |
| Current smoker | 10257 (100.0) | 1039 (100.0) |
| eGFR | 9784 (95.4) | 1038 (99.9) |
| Age at diabetes diagnosis | 9908 (96.6) | 626 (60.3) |
| HbA_1c_ | 9737 (94.9) | 1034 (99.5) |
| Total-C | 10257 (100.0) | 1039 (100.0) |
| HDL-C | 10257 (100.0) | 1039 (100.0) |
| non-HDL-C | 10257 (100.0) | 1039 (100.0) |
| Remnant-C | 10257 (100.0) | 1039 (100.0) |
| VLDL-C | 10257 (100.0) | 1039 (100.0) |
| Clinical-LDL-C | 10257 (100.0) | 1039 (100.0) |
| LDL-C | 10257 (100.0) | 1039 (100.0) |
| Total-TG | 10257 (100.0) | 1039 (100.0) |
| VLDL-TG | 10257 (100.0) | 1039 (100.0) |
| LDL-TG | 10257 (100.0) | 1039 (100.0) |
| HDL-TG | 10257 (100.0) | 1039 (100.0) |
| Total-PL | 10257 (100.0) | 1039 (100.0) |
| VLDL-PL | 10257 (100.0) | 1039 (100.0) |
| LDL-PL | 10257 (100.0) | 1039 (100.0) |
| HDL-PL | 10257 (100.0) | 1039 (100.0) |
| Total-CE | 10257 (100.0) | 1039 (100.0) |
| VLDL-CE | 10257 (100.0) | 1039 (100.0) |
| LDL-CE | 10257 (100.0) | 1039 (100.0) |
| HDL-CE | 10257 (100.0) | 1039 (100.0) |
| Total-FC | 10257 (100.0) | 1039 (100.0) |
| VLDL-FC | 10257 (100.0) | 1039 (100.0) |
| LDL-FC | 10257 (100.0) | 1039 (100.0) |
| HDL-FC | 10257 (100.0) | 1039 (100.0) |
| Total-L | 10257 (100.0) | 1039 (100.0) |
| VLDL-L | 10257 (100.0) | 1039 (100.0) |
| LDL-L | 10257 (100.0) | 1039 (100.0) |
| HDL-L | 10257 (100.0) | 1039 (100.0) |
| Total-P | 10257 (100.0) | 1039 (100.0) |
| VLDL-P | 10257 (100.0) | 1039 (100.0) |
| LDL-P | 10257 (100.0) | 1039 (100.0) |
| HDL-P | 10257 (100.0) | 1039 (100.0) |
| VLDL-size | 10257 (100.0) | 1039 (100.0) |
| LDL-size | 10257 (100.0) | 1039 (100.0) |
| HDL-size | 10257 (100.0) | 1039 (100.0) |
| Phosphoglyc | 10247 (99.9) | 1035 (99.6) |
| TG-by-PG | 10247 (99.9) | 1035 (99.6) |
| Cholines | 10247 (99.9) | 1035 (99.6) |
| Phosphatidylc | 10247 (99.9) | 1035 (99.6) |
| Sphingomyelins | 10247 (99.9) | 1035 (99.6) |
| ApoB | 10257 (100.0) | 1039 (100.0) |
| ApoA1 | 10257 (100.0) | 1039 (100.0) |
| ApoB-by-ApoA1 | 10257 (100.0) | 1039 (100.0) |
| Total-FA | 10247 (99.9) | 1035 (99.6) |
| Unsaturation | 10247 (99.9) | 1035 (99.6) |
| Omega-3 | 10247 (99.9) | 1035 (99.6) |
| Omega-6 | 10247 (99.9) | 1035 (99.6) |
| PUFA | 10247 (99.9) | 1035 (99.6) |
| MUFA | 10247 (99.9) | 1035 (99.6) |
| SFA | 10247 (99.9) | 1035 (99.6) |
| LA | 10247 (99.9) | 1035 (99.6) |
| DHA | 10247 (99.9) | 1035 (99.6) |
| Omega-3-pct | 10247 (99.9) | 1035 (99.6) |
| Omega-6-pct | 10247 (99.9) | 1035 (99.6) |
| PUFA-pct | 10247 (99.9) | 1035 (99.6) |
| MUFA-pct | 10247 (99.9) | 1035 (99.6) |
| SFA-pct | 10247 (99.9) | 1035 (99.6) |
| LA-pct | 10247 (99.9) | 1035 (99.6) |
| DHA-pct | 10247 (99.9) | 1035 (99.6) |
| PUFA-by-MUFA | 10247 (99.9) | 1035 (99.6) |
| Omega-6-by-Omega-3 | 10247 (99.9) | 1035 (99.6) |
| Ala | 10255 (100.0) | 1032 (99.3) |
| Gln | 10205 (99.5) | 997 (96.0) |
| Gly | 10197 (99.4) | 1034 (99.5) |
| His | 10240 (99.8) | 1034 (99.5) |
| Total-BCAA | 10249 (99.9) | 1036 (99.7) |
| Ile | 10257 (100.0) | 1039 (100.0) |
| Leu | 10257 (100.0) | 1039 (100.0) |
| Val | 10249 (99.9) | 1036 (99.7) |
| Phe | 10245 (99.9) | 1034 (99.5) |
| Tyr | 10245 (99.9) | 1034 (99.5) |
| Glucose | 10239 (99.8) | 938 (90.3) |
| Lactate | 10240 (99.8) | 1038 (99.9) |
| Pyruvate | 10206 (99.5) | 1008 (97.0) |
| Citrate | 10256 (100.0) | 1011 (97.3) |
| bOHbutyrate | 10076 (98.2) | 1036 (99.7) |
| Acetate | 10246 (99.9) | 1039 (100.0) |
| Acetoacetate | 10257 (100.0) | 1038 (99.9) |
| Acetone | 10257 (100.0) | 1039 (100.0) |
| Creatinine | 10029 (97.8) | 977 (94.0) |
| Albumin | 10252 (100.0) | 1039 (100.0) |
| GlycA | 10257 (100.0) | 1039 (100.0) |
| XXL-VLDL-P | 10257 (100.0) | 1039 (100.0) |
| XXL-VLDL-L | 10257 (100.0) | 1039 (100.0) |
| XXL-VLDL-PL | 10257 (100.0) | 1039 (100.0) |
| XXL-VLDL-C | 10257 (100.0) | 1039 (100.0) |
| XXL-VLDL-CE | 10257 (100.0) | 1039 (100.0) |
| XXL-VLDL-FC | 10257 (100.0) | 1039 (100.0) |
| XXL-VLDL-TG | 10257 (100.0) | 1039 (100.0) |
| XL-VLDL-P | 10257 (100.0) | 1039 (100.0) |
| XL-VLDL-L | 10257 (100.0) | 1039 (100.0) |
| XL-VLDL-PL | 10257 (100.0) | 1039 (100.0) |
| XL-VLDL-C | 10257 (100.0) | 1039 (100.0) |
| XL-VLDL-CE | 10257 (100.0) | 1039 (100.0) |
| XL-VLDL-FC | 10257 (100.0) | 1039 (100.0) |
| XL-VLDL-TG | 10257 (100.0) | 1039 (100.0) |
| L-VLDL-P | 10257 (100.0) | 1039 (100.0) |
| L-VLDL-L | 10257 (100.0) | 1039 (100.0) |
| L-VLDL-PL | 10257 (100.0) | 1039 (100.0) |
| L-VLDL-C | 10257 (100.0) | 1039 (100.0) |
| L-VLDL-CE | 10257 (100.0) | 1039 (100.0) |
| L-VLDL-FC | 10257 (100.0) | 1039 (100.0) |
| L-VLDL-TG | 10257 (100.0) | 1039 (100.0) |
| M-VLDL-P | 10257 (100.0) | 1039 (100.0) |
| M-VLDL-L | 10257 (100.0) | 1039 (100.0) |
| M-VLDL-PL | 10257 (100.0) | 1039 (100.0) |
| M-VLDL-C | 10257 (100.0) | 1039 (100.0) |
| M-VLDL-CE | 10257 (100.0) | 1039 (100.0) |
| M-VLDL-FC | 10257 (100.0) | 1039 (100.0) |
| M-VLDL-TG | 10257 (100.0) | 1039 (100.0) |
| S-VLDL-P | 10257 (100.0) | 1039 (100.0) |
| S-VLDL-L | 10257 (100.0) | 1039 (100.0) |
| S-VLDL-PL | 10257 (100.0) | 1039 (100.0) |
| S-VLDL-C | 10257 (100.0) | 1039 (100.0) |
| S-VLDL-CE | 10257 (100.0) | 1039 (100.0) |
| S-VLDL-FC | 10257 (100.0) | 1039 (100.0) |
| S-VLDL-TG | 10257 (100.0) | 1039 (100.0) |
| XS-VLDL-P | 10257 (100.0) | 1039 (100.0) |
| XS-VLDL-L | 10257 (100.0) | 1039 (100.0) |
| XS-VLDL-PL | 10257 (100.0) | 1039 (100.0) |
| XS-VLDL-C | 10257 (100.0) | 1039 (100.0) |
| XS-VLDL-CE | 10257 (100.0) | 1039 (100.0) |
| XS-VLDL-FC | 10257 (100.0) | 1039 (100.0) |
| XS-VLDL-TG | 10257 (100.0) | 1039 (100.0) |
| IDL-P | 10257 (100.0) | 1039 (100.0) |
| IDL-L | 10257 (100.0) | 1039 (100.0) |
| IDL-PL | 10257 (100.0) | 1039 (100.0) |
| IDL-C | 10257 (100.0) | 1039 (100.0) |
| IDL-CE | 10257 (100.0) | 1039 (100.0) |
| IDL-FC | 10257 (100.0) | 1039 (100.0) |
| IDL-TG | 10257 (100.0) | 1039 (100.0) |
| L-LDL-P | 10257 (100.0) | 1039 (100.0) |
| L-LDL-L | 10257 (100.0) | 1039 (100.0) |
| L-LDL-PL | 10257 (100.0) | 1039 (100.0) |
| L-LDL-C | 10257 (100.0) | 1039 (100.0) |
| L-LDL-CE | 10257 (100.0) | 1039 (100.0) |
| L-LDL-FC | 10257 (100.0) | 1039 (100.0) |
| L-LDL-TG | 10257 (100.0) | 1039 (100.0) |
| M-LDL-P | 10257 (100.0) | 1039 (100.0) |
| M-LDL-L | 10257 (100.0) | 1039 (100.0) |
| M-LDL-PL | 10257 (100.0) | 1039 (100.0) |
| M-LDL-C | 10257 (100.0) | 1039 (100.0) |
| M-LDL-CE | 10257 (100.0) | 1039 (100.0) |
| M-LDL-FC | 10257 (100.0) | 1039 (100.0) |
| M-LDL-TG | 10257 (100.0) | 1039 (100.0) |
| S-LDL-P | 10257 (100.0) | 1039 (100.0) |
| S-LDL-L | 10257 (100.0) | 1039 (100.0) |
| S-LDL-PL | 10257 (100.0) | 1039 (100.0) |
| S-LDL-C | 10257 (100.0) | 1039 (100.0) |
| S-LDL-CE | 10257 (100.0) | 1039 (100.0) |
| S-LDL-FC | 10257 (100.0) | 1039 (100.0) |
| S-LDL-TG | 10257 (100.0) | 1039 (100.0) |
| XL-HDL-P | 10257 (100.0) | 1039 (100.0) |
| XL-HDL-L | 10257 (100.0) | 1039 (100.0) |
| XL-HDL-PL | 10257 (100.0) | 1039 (100.0) |
| XL-HDL-C | 10257 (100.0) | 1039 (100.0) |
| XL-HDL-CE | 10257 (100.0) | 1039 (100.0) |
| XL-HDL-FC | 10257 (100.0) | 1039 (100.0) |
| XL-HDL-TG | 10257 (100.0) | 1039 (100.0) |
| L-HDL-P | 10257 (100.0) | 1039 (100.0) |
| L-HDL-L | 10257 (100.0) | 1039 (100.0) |
| L-HDL-PL | 10257 (100.0) | 1039 (100.0) |
| L-HDL-C | 10257 (100.0) | 1039 (100.0) |
| L-HDL-CE | 10257 (100.0) | 1039 (100.0) |
| L-HDL-FC | 10257 (100.0) | 1039 (100.0) |
| L-HDL-TG | 10257 (100.0) | 1039 (100.0) |
| M-HDL-P | 10257 (100.0) | 1039 (100.0) |
| M-HDL-L | 10257 (100.0) | 1039 (100.0) |
| M-HDL-PL | 10257 (100.0) | 1039 (100.0) |
| M-HDL-C | 10257 (100.0) | 1039 (100.0) |
| M-HDL-CE | 10257 (100.0) | 1039 (100.0) |
| M-HDL-FC | 10257 (100.0) | 1039 (100.0) |
| M-HDL-TG | 10257 (100.0) | 1039 (100.0) |
| S-HDL-P | 10257 (100.0) | 1039 (100.0) |
| S-HDL-L | 10257 (100.0) | 1039 (100.0) |
| S-HDL-PL | 10257 (100.0) | 1039 (100.0) |
| S-HDL-C | 10257 (100.0) | 1039 (100.0) |
| S-HDL-CE | 10257 (100.0) | 1039 (100.0) |
| S-HDL-FC | 10257 (100.0) | 1039 (100.0) |
| S-HDL-TG | 10257 (100.0) | 1039 (100.0) |
| XXL-VLDL-PL-pct | 10257 (100.0) | 1039 (100.0) |
| XXL-VLDL-C-pct | 10257 (100.0) | 1039 (100.0) |
| XXL-VLDL-CE-pct | 10257 (100.0) | 1039 (100.0) |
| XXL-VLDL-FC-pct | 10257 (100.0) | 1039 (100.0) |
| XXL-VLDL-TG-pct | 10257 (100.0) | 1039 (100.0) |
| XL-VLDL-PL-pct | 10257 (100.0) | 1039 (100.0) |
| XL-VLDL-C-pct | 10257 (100.0) | 1039 (100.0) |
| XL-VLDL-CE-pct | 10257 (100.0) | 1039 (100.0) |
| XL-VLDL-FC-pct | 10257 (100.0) | 1039 (100.0) |
| XL-VLDL-TG-pct | 10257 (100.0) | 1039 (100.0) |
| L-VLDL-PL-pct | 10257 (100.0) | 1039 (100.0) |
| L-VLDL-C-pct | 10257 (100.0) | 1039 (100.0) |
| L-VLDL-CE-pct | 10257 (100.0) | 1039 (100.0) |
| L-VLDL-FC-pct | 10257 (100.0) | 1039 (100.0) |
| L-VLDL-TG-pct | 10257 (100.0) | 1039 (100.0) |
| M-VLDL-PL-pct | 10257 (100.0) | 1039 (100.0) |
| M-VLDL-C-pct | 10257 (100.0) | 1039 (100.0) |
| M-VLDL-CE-pct | 10257 (100.0) | 1039 (100.0) |
| M-VLDL-FC-pct | 10257 (100.0) | 1039 (100.0) |
| M-VLDL-TG-pct | 10257 (100.0) | 1039 (100.0) |
| S-VLDL-PL-pct | 10257 (100.0) | 1039 (100.0) |
| S-VLDL-C-pct | 10257 (100.0) | 1039 (100.0) |
| S-VLDL-CE-pct | 10257 (100.0) | 1039 (100.0) |
| S-VLDL-FC-pct | 10257 (100.0) | 1039 (100.0) |
| S-VLDL-TG-pct | 10257 (100.0) | 1039 (100.0) |
| XS-VLDL-PL-pct | 10257 (100.0) | 1039 (100.0) |
| XS-VLDL-C-pct | 10257 (100.0) | 1039 (100.0) |
| XS-VLDL-CE-pct | 10257 (100.0) | 1039 (100.0) |
| XS-VLDL-FC-pct | 10257 (100.0) | 1039 (100.0) |
| XS-VLDL-TG-pct | 10257 (100.0) | 1039 (100.0) |
| IDL-PL-pct | 10257 (100.0) | 1039 (100.0) |
| IDL-C-pct | 10257 (100.0) | 1039 (100.0) |
| IDL-CE-pct | 10257 (100.0) | 1039 (100.0) |
| IDL-FC-pct | 10257 (100.0) | 1039 (100.0) |
| IDL-TG-pct | 10257 (100.0) | 1039 (100.0) |
| L-LDL-PL-pct | 10257 (100.0) | 1039 (100.0) |
| L-LDL-C-pct | 10257 (100.0) | 1039 (100.0) |
| L-LDL-CE-pct | 10257 (100.0) | 1039 (100.0) |
| L-LDL-FC-pct | 10257 (100.0) | 1039 (100.0) |
| L-LDL-TG-pct | 10257 (100.0) | 1039 (100.0) |
| M-LDL-PL-pct | 10257 (100.0) | 1039 (100.0) |
| M-LDL-C-pct | 10257 (100.0) | 1039 (100.0) |
| M-LDL-CE-pct | 10257 (100.0) | 1039 (100.0) |
| M-LDL-FC-pct | 10257 (100.0) | 1039 (100.0) |
| M-LDL-TG-pct | 10257 (100.0) | 1039 (100.0) |
| S-LDL-PL-pct | 10257 (100.0) | 1039 (100.0) |
| S-LDL-C-pct | 10257 (100.0) | 1039 (100.0) |
| S-LDL-CE-pct | 10257 (100.0) | 1039 (100.0) |
| S-LDL-FC-pct | 10257 (100.0) | 1039 (100.0) |
| S-LDL-TG-pct | 10257 (100.0) | 1039 (100.0) |
| XL-HDL-PL-pct | 10257 (100.0) | 1039 (100.0) |
| XL-HDL-C-pct | 10257 (100.0) | 1039 (100.0) |
| XL-HDL-CE-pct | 10257 (100.0) | 1039 (100.0) |
| XL-HDL-FC-pct | 10257 (100.0) | 1039 (100.0) |
| XL-HDL-TG-pct | 10257 (100.0) | 1039 (100.0) |
| L-HDL-PL-pct | 10257 (100.0) | 1039 (100.0) |
| L-HDL-C-pct | 10257 (100.0) | 1039 (100.0) |
| L-HDL-CE-pct | 10257 (100.0) | 1039 (100.0) |
| L-HDL-FC-pct | 10257 (100.0) | 1039 (100.0) |
| L-HDL-TG-pct | 10257 (100.0) | 1039 (100.0) |
| M-HDL-PL-pct | 10257 (100.0) | 1039 (100.0) |
| M-HDL-C-pct | 10257 (100.0) | 1039 (100.0) |
| M-HDL-CE-pct | 10257 (100.0) | 1039 (100.0) |
| M-HDL-FC-pct | 10257 (100.0) | 1039 (100.0) |
| M-HDL-TG-pct | 10257 (100.0) | 1039 (100.0) |
| S-HDL-PL-pct | 10257 (100.0) | 1039 (100.0) |
| S-HDL-C-pct | 10257 (100.0) | 1039 (100.0) |
| S-HDL-CE-pct | 10257 (100.0) | 1039 (100.0) |
| S-HDL-FC-pct | 10257 (100.0) | 1039 (100.0) |
| S-HDL-TG-pct | 10257 (100.0) | 1039 (100.0) |

**Abbreviations:** Ala, alanine; Apo-A1=apolipoprotein A1; Apo-B=apolipoprotein B; bOHbutyrate, 3-Hydroxybutyrate; BCAA= Branched-Chain Amino Acids; C=cholesterol; CE=cholesteryl esters; DHA=docosahexaenoic acid; eGFR, estimated Glomerular Filtration Rate; FA=fatty acids; Gln, glutamine; FC= free cholesterol; Gly, glycine; GlycA, glycoprotein acetyls; HbA_1c_, glycated hemoglobin; HDL=high density lipoproteins; HDL-D=high density lipoprotein particle diameter; His, histidine; IDL=intermediate density lipoproteins; Ile, isoleucine; L=large; LA=linoleic acid; LDL=low density lipoproteins; LDL-D=low density lipoprotein particle diameter; Leu, leucine; LP=lipoprotein; M=medium; MUFA=monounsaturated fatty acids; P=particles; Phe, phenylalanine; PL=Phospholipids; PUFA=polyunsaturated fatty acids; pct= percentage; S=small; SFA=saturated fatty acids; TG=triglycerides; Tyr, tyrosine; Val, valine; VLDL=very low density lipoproteins; VLDL-D=very low density lipoprotein particle diameter; XL=very large; XS=very small; XXL=extremely large.

**Table S2.** Laboratory methods applied in the UK Biobank (UKB) and ESTHER study

| **Biomarker** | **Study** | **Analytical method** | **Instrument** | **Supplier** |
| --- | --- | --- | --- | --- |
| HDL-C | UKB | Enzymatic | Beckman Coulter AU5800 | Beckman Coulter (UK), Ltd |
| HDL-C | ESTHER | Enzymatic | Cobas 8000 C701 | Roche Diagnostics, Mannheim, Germany |
| Total cholesterol | UKB | Enzymatic | Beckman Coulter AU5800 | Beckman Coulter (UK), Ltd |
| Total cholesterol | ESTHER | Enzymatic | Cobas 8000 C701 | Roche Diagnostics, Mannheim, Germany |
| HbA_1c_ | UKB | HPLC | Bio-Rad Variant II | Bio-Rad Laboratories, Hercules, CA, USA |
| HbA_1c_ | ESTHER | HPLC | Bio-Rad Variant II | Bio-Rad Laboratories, Hercules, CA, USA |
| Creatinine | UKB | Enzymatic | Beckman Coulter AU5800 | Beckman Coulter (UK), Ltd |
| Creatinine | ESTHER | Kinetic Jaffé method | Cobas 8000 C701 | Roche Diagnostics, Mannheim, Germany |

**Abbreviations:** HbA_1c_, glycated hemoglobin; HDL-C, high-density lipoprotein cholesterol; HPLC, high pressure liquid chromatography.

**Table S3.** Definition of endpoint major cardiovascular event (MACE)

| **Fatal MACE – cause-specific mortality due to any of the following:** | |
| --- | --- |
| *Endpoints included* | *ICD10-codes* |
| Hypertensive disease | I10-16 |
| Ischemic heart disease | I20-25 |
| Arrhythmias, heart failure | I46-52 |
| Cerebrovascular disease | I60-69 |
| Atherosclerosis/aortic aneurysm | I70-73 |
| Sudden death and death within 24 hours of symptom onset | R96.0-96.1 |
| *Endpoints excluded from the above endpoint:* | |
| Myocarditis, unspecified | I51.4 |
| Subarachnoid haemorrhage | I60 |
| Subdural hemorrhage | I62 |
| Cerebral aneurysm | I67.1 |
| Cerebral arteritis | I68.2 |
| Moyamoya | I67.5 |
| **Non-fatal MACE** | |
| Non-fatal myocardial infarction | I21-I23 |
| Non-fatal stroke | I60-69* |

***** In UK Biobank: I61, I63-I66, I69; in ESTHER: I60-I69

### Table S4. ß-coefficients of the variables of the SCORE2-Diabetes model extended by 7 metabolites for 10-year prediction of major cardiovascular events in the derivation set of the UK Biobank

| Risk factor (units) | ß-coefficient | |
| --- | --- | --- |
|  | Men | Women |
| **SCORE2-Diabetes variables** |  |  |
| Age (per 5 years) | 0.3649 | 0.4972 |
| Current smoking | 0.1815 | 0.7378 |
| Systolic blood pressure (per 20 mmHg) | 0.0831 | 0.2404 |
| Total cholesterol (per 1 mmol/L) | 0.0060 | 0.1019 |
| HDL cholesterol (per 0.5 mmol/L) | 0.2256 | -0.1160 |
| Smoking interaction with age | -0.2768 | -0.1317 |
| SBP interaction with age | -0.0260 | -0.0815 |
| Total cholesterol interaction with age | -0.0363 | -0.0102 |
| HDL interaction with age | 0.0509 | -0.0408 |
| Diabetes age at diagnosis (per 5 years) | -0.1146 | -0.1317 |
| HbA_1c_ (per 9.34 mmol/mol) | 0.1368 | 0.1098 |
| Ln eGFR (per 0.15 ml/min/1.73m^2^) | -0.1393 | -0.0947 |
| Ln eGFR^2^ (quadratic term) | -0.0067 | -0.0051 |
| HbA_1c_ interaction with age | 0.0363 | 0.0069 |
| Ln eGFR interaction with age | 0.0182 | -0.0208 |
| Diabetes | 0.6457 | 0.8096 |
| **Added metabolites** |  |  |
| Creatinine (per 1 SD^a^) | 0.0341 | 0.0652 |
| Albumin (per 1 SD^a^) | -0.0491 | -0.2865 |
| GlycA (per 1 SD^a^) | 0.1704 | 0.1283 |
| Acetate (per 1 SD^a^) | 0.0395 | - |
| Omega-3-pct (per 1 SD^a^) | -0.0682 | - |
| VLDL-size (per 1 SD^a^) | -0.1076 | - |
| Lactate (per 1 SD^a^) | - | 0.1282 |

**Abbreviations:** CI, confidence interval; GlycA, glycoprotein acetyls; HbA_1c_, glycated hemoglobin; SHR, subdistribution hazard ratio; Omega-3-pct, the Omega-3 fatty acids percentage of total fatty acids; SD, standard deviation; VLDL-size, average diameter for very-low-density lipoprotein particles.

^a^ The standard deviations of the metabolites creatinine, albumin, GlycA, acetate, omega-3-pct, VLDL-size, and lactate were 0.02 mmol/L, 3.60 mmol/L, 0.13 mmol/L, 0.04 mmol/L, 1.51%, 1.41 mmol/L, and 1.23 mmol/L, respectively.

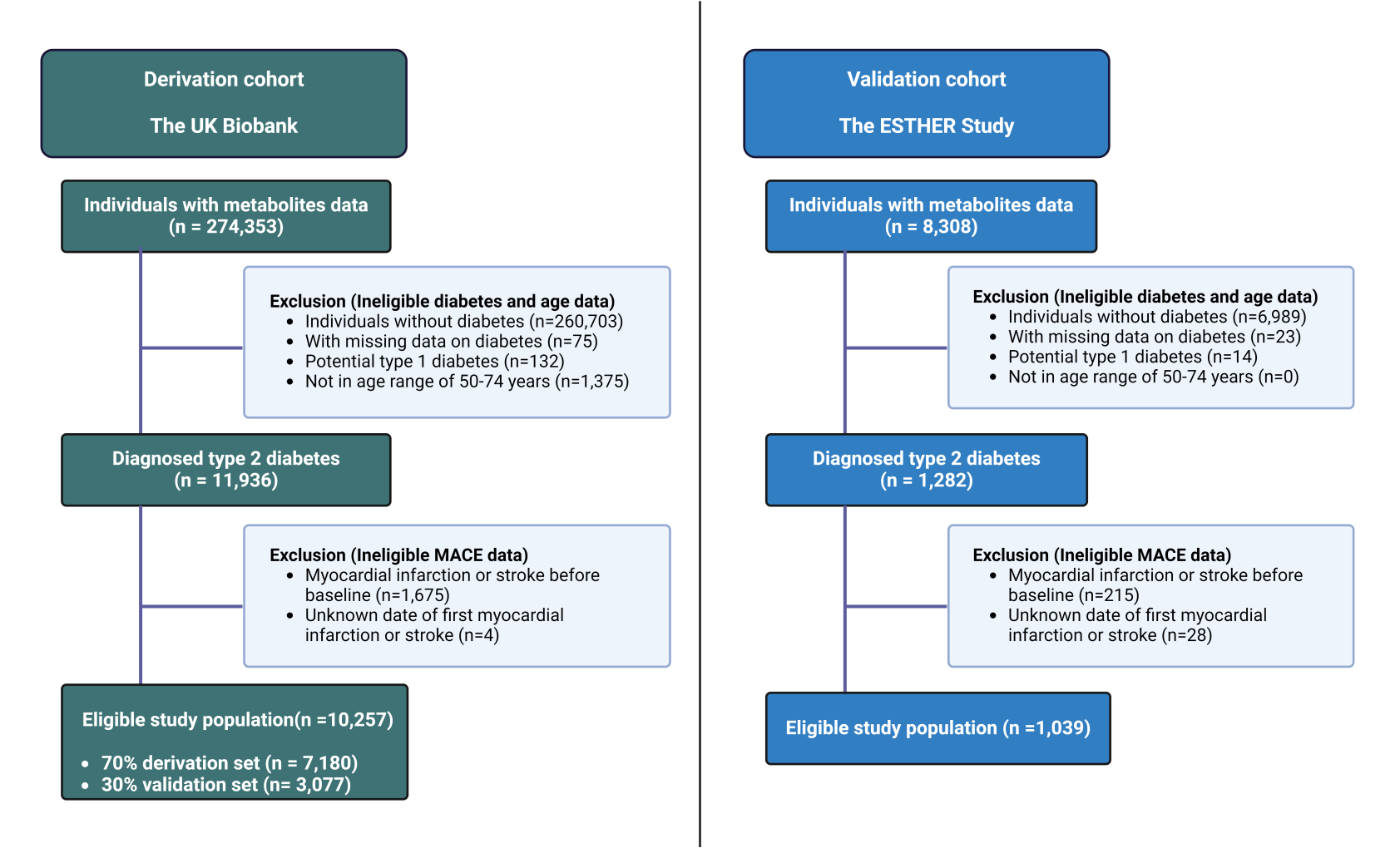

### Figure S1. Flow chart for participant inclusion and exclusion

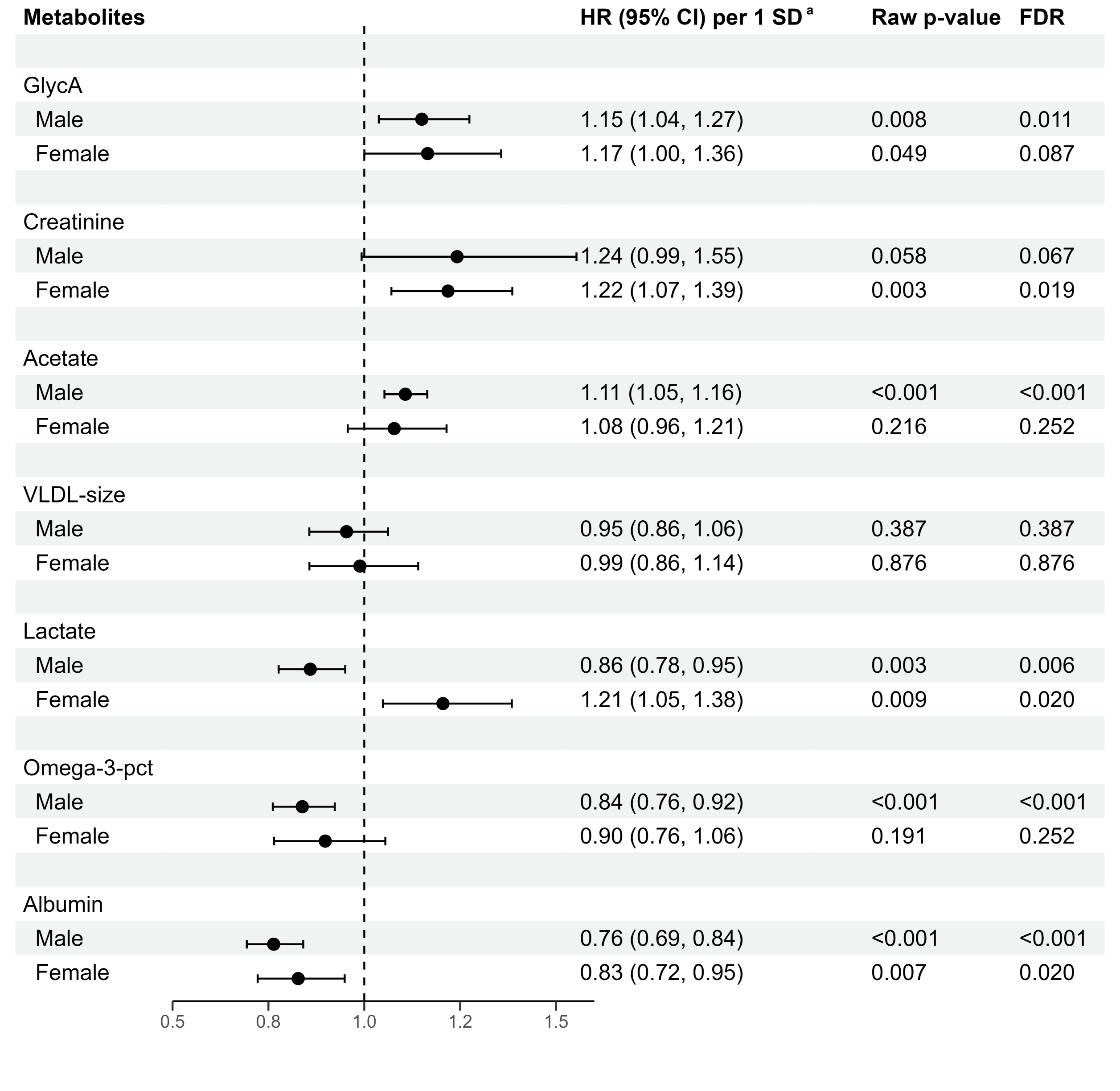

**Figure S2.** Associations between selected metabolites and major cardiovascular events across sexes in the internal validation cohort (30% of UK Biobank, N=3,077)

**Abbreviations:** CI, confidence interval; FDR, false discovery rate; GlycA, glycoprotein acetyls; HR, hazard ratio; Omega-3-pct, the Omega-3 fatty acids percentage of total fatty acids; SD, standard deviation; VLDL-size, average diameter for very-low-density lipoprotein particles.

**^a^** Hazard ratios are expressed per 1 standard deviation of the respective metabolite concentration and are adjusted for age, systolic blood pressure, smoking status, diabetes age at diagnosis, glycated hemoglobin, and the estimated glomerular filtration rate. The standard deviations of creatinine, albumin, GlycA, acetate, omega-3-pct, VLDL-size, and lactate were 0.02 mmol/L, 3.60 mmol/L, 0.13 mmol/L, 0.04 mmol/L, 1.51%, 1.41 mmol/L, and 1.23 mmol/L, respectively.

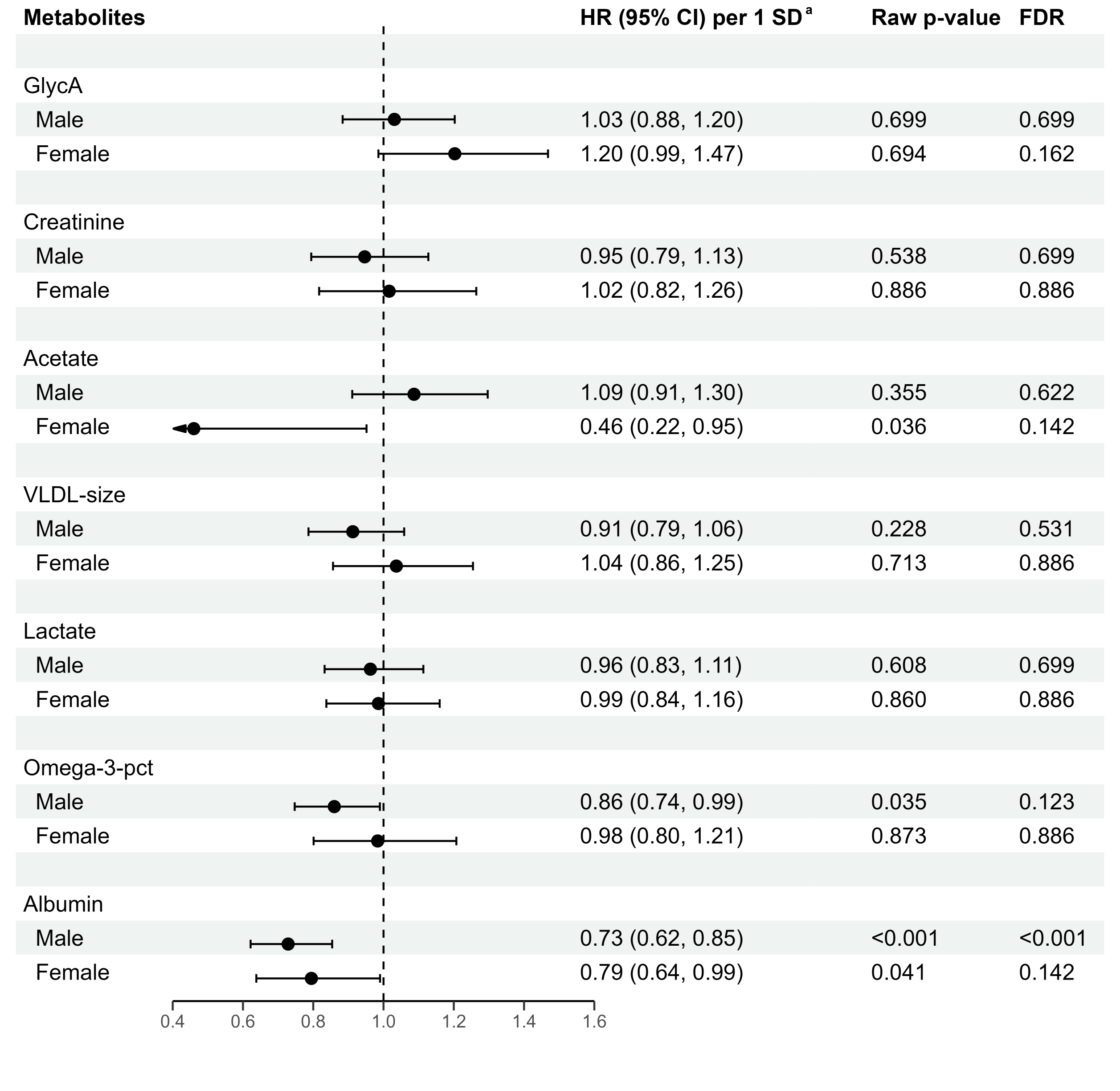

### Figure S3. Associations between selected metabolites and major cardiovascular events across sexes in the external validation cohort (ESTHER study, N=1,039)

**Abbreviations:** CI, confidence interval; GlycA, glycoprotein acetyls; HR, hazard ratio; Omega-3-pct, the Omega-3 fatty acids percentage of total fatty acids; VLDL-size, average diameter for very-low-density lipoprotein particles.

**^a^** Hazard ratios are expressed per 1 standard deviation of the respective metabolite concentration and are adjusted for age, systolic blood pressure, smoking status, diabetes age at diagnosis, glycated hemoglobin, and estimated glomerular filtration rate. The standard deviations of creatinine, albumin, GlycA, acetate, omega-3-pct, VLDL-size, and lactate were 0.03 mmol/L, 4.08 mmol/L, 0.14 mmol/L, 0.04 mmol/L, 1.19%, 1.08 mmol/L, and 5.36 mmol/L, respectively.

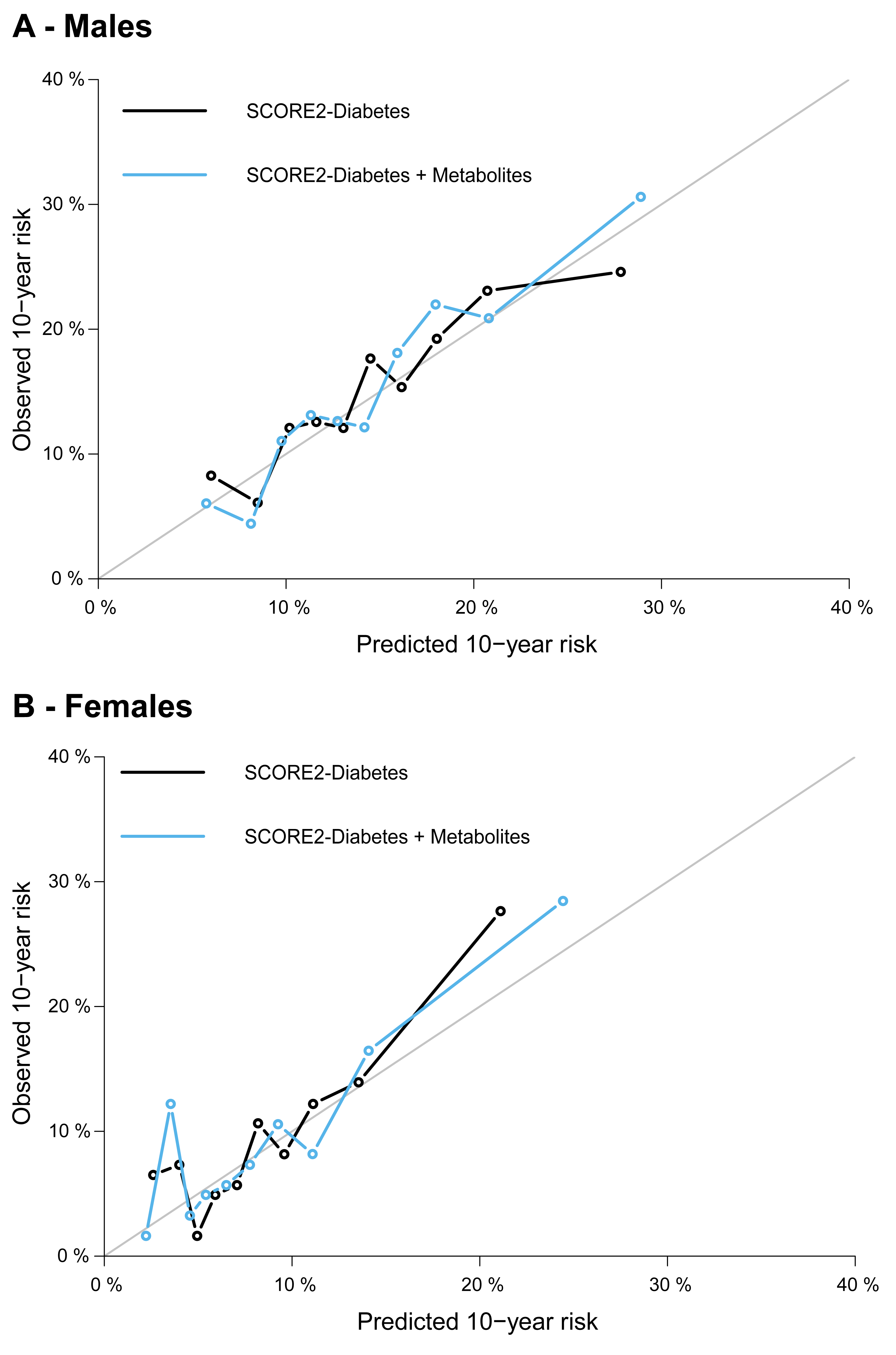

### Figure S4. Calibration curves of the SCORE2-Diabetes model and its combination with the metabolites for 10-year cardiovascular risk prediction in the internal validation set (30% of UK Biobank, N=3,077)

**Note:** The metabolomic biomarkers added were creatinine, albumin, GlycA, acetate, Omega-3-pct, VLDL-size for male, and lactate, creatinine, albumin, GlycA for female (see **Table S4** for abbreviations).

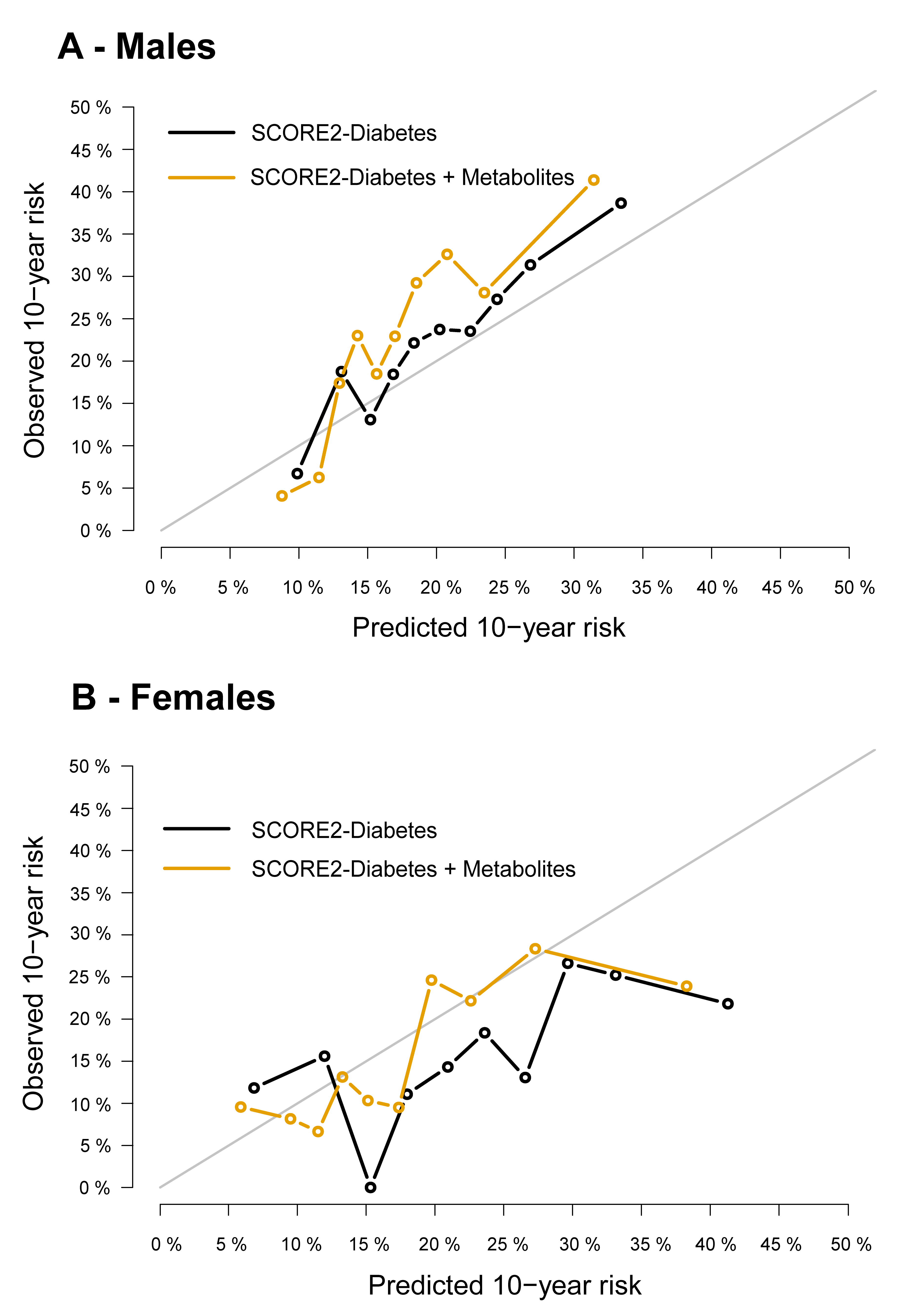

### Figure S5. Calibration curves of the SCORE2-Diabetes model and its combination with the metabolites for 10-year cardiovascular risk prediction in the external validation (ESTHER study, N=1,039)

**Note:** The metabolomic biomarkers added were creatinine, albumin, GlycA, acetate, Omega-3-pct, VLDL-size for male, and lactate, creatinine, albumin, GlycA for female (see **Table S4** for abbreviations).
